# Communicating Ethically Relevant Practices of Patient Registries: An Interview Study on the Information Needs of People with Multiple Sclerosis

**DOI:** 10.64898/2026.09.05.26362329

**Authors:** Olmo R. van den Akker, Fabian Windfuhr, Diogo Almeida, Sieta T. de Vries, Susanne Stark

## Abstract

Patient registries play a key role in research on chronic conditions like multiple sclerosis (MS), but their success typically depends on patients’ willingness to share data. Information provided about ethically relevant registry practices may strongly influence their willingness. This study explores what information about ethics practices people with MS consider important when deciding whether to contribute data to a patient registry, and how they prefer this information to be communicated. We conducted semi-structured online interviews with 21 people with MS and patient representatives from nine European countries. The interview guide first addressed general information needs and then seven predefined ethically relevant topics. The topics registry governance, registry funding, data handling, and use-and-access procedures were adapted from the literature, while communication, access, and rewards for contributing data provided contextual and practical insight. They also served as the initial coding categories for directed qualitative content analysis, after which categories were refined and reorganized during analysis, and subcategories were inductively developed. We found that trust emerged as a cross-cutting factor. Participants required less information about registry practices when they trusted the registry or associated parties and preferred registry communication through trusted intermediaries. When deciding whether to contribute data to a patient registry, participants emphasized the importance of clarity about how the data will be used, who would have access to it, and what privacy protections are in place, while information preferences for technical details regarding data use and access varied widely. Feedback on personal or aggregate results was highly valued and seen as types of personal and communal benefits for participating. In conclusion, patient registries should adopt flexible communication strategies that balance transparency and accessibility. Building and maintaining trust, potentially through trusted affiliates like universities or patient organizations, appears central to properly inform (potential) registry participants about ethically relevant registry practices.

## Introduction

Patient registries are valuable tools in advancing research and improving care for chronic conditions such as multiple sclerosis (MS; Flachenecker and Stuke, 2008; Hillert and Stawiarz, 2015). They have been established worldwide to systematically collect data on diagnoses, sociodemographic variables, medical treatment histories, or care pathways (Hageman et al., 2023). An example in MS is the Swedish MS registry, SMSreg, covering about 80% of the MS population in Sweden since 2001 (Hillert and Stawiarz, 2015). The data from registries add value to other types of data (e.g., from clinical trials) because they often cover larger populations over a longer time span (Agency for Healthcare Research and Quality, 2020; Jonker et al., 2022). Patient registries therefore allow monitoring the long-term health status of individual patients and can provide deeper insights into a disease. The success of registries depends on how they collect, curate, and document data, and on how they report any findings (Agency for Healthcare Research and Quality, 2020), though patients sharing their data is a fundamental prerequisite. Studies show that there are several factors that influence patients’ decisions to share data. Among them are patients’ attitudes toward the registry’s ethically relevant practices and their perception of being adequately informed about these practices (Baines et al., 2024; Kalkman et al., 2022; Simon et al., 2009).

Especially in the case of complex, lifelong conditions like MS, that often require sustained engagement with the healthcare system, deciding whether to contribute health data to a registry involves significant ethical considerations (Baird et al., 2009). For example, concerns about who will have access to one’s data and how one’s privacy is protected can play a crucial role in shaping decisions to share data (Cascini et al., 2024; Kalkman et al., 2022). Registries should therefore be transparent toward the public about how they operate. While the importance of transparency is recognized in institutional documents like the guide for good registry practices from the Agency for Healthcare Research and Quality (2020) and the guideline on registry-based studies from the European Medicines Agency (2021), there remains a gap in understanding what specific information people want to receive in order to feel comfortable about their participation in a registry, and how they would want to receive this information. This is an important gap to fill because earlier studies show that the information provided by registries is not always optimal (Van den Akker et al., 2024; Whiddett et al., 2006). As such, the research questions (RQs) in this study are:

1. **“What information about ethically relevant registry practices do people with MS consider important when deciding whether to contribute their health data to a patient registry?”**
2. **“How do people with MS want information about these registry practices to be communicated?”**

The knowledge gleaned from this study can inform registries’ practices and communication strategies so that they are responsive to patients’ values and expectations.

## Methods

### Study design and population

We conducted a qualitative study using semi-structured online interviews in English among people with MS and MS patient representatives living in Europe. We targeted people with MS for their individual perspectives and patient representatives because they are in contact with many individual patients and would therefore be able to provide a broad perspective of the MS community.

A convenience sample of potential participants was invited for interviews in three waves by the European Multiple Sclerosis Platform (EMSP). In the first wave, the EMSP sent an invitation email with general information about the study to its members, mentioned the study invitation in its newsletter, and shared it via their social media channels. In the second wave, EMSP sent individual email reminders to their members in specific countries (i.e., Norway, Spain, Portugal, Italy, and Germany) to increase the breadth of our included sample. In the final wave, EMSP’s Community Advisory Board members from Spain and Portugal were contacted specifically to encourage their participation. Participants were offered an Amazon voucher worth 25 euro for participating.

### Data collection

Participants could email the study team to express their interest. We then emailed them back and invited them to read an information letter (see https://osf.io/qakzr), give their consent for participation, and fill out a short survey using the REDcap platform (see https://osf.io/heqmz) asking about demographics, whether they had MS and, if so, for how long, whether they acted as a patient representative, and whether they were currently or ever enrolled in a registry. Once they submitted their responses, we followed up with an email to schedule a meeting (see https://osf.io/94msv).

The interviews were semi-structured and consisted of two main parts: one that focused on the information that participants want to receive from MS registries and how, and one that focused on patient-reported outcomes that participants could share with MS registries. This paper reports on the first part of the interviews. Interviews were conducted digitally via Microsoft Teams.

We first asked participants about their general information needs when it comes to registry participation, and later about their information needs regarding five ethics topics that were taken from Van den Akker et al. (2024): governance, conflicts of interest, informed consent, privacy and data protection, and use-and-access procedures. Van den Akker et al. developed this list of registry ethics practices based on two popular assessment tools for patient registries: the Registry Evaluation and Quality Standards Tool, REQueST (EUnetHTA, 2019), and the Agency for Healthcare Research and Quality’s (2020) guide for good registry practice, and a systematic review on the ethics of data sharing (Kalkman et al., 2019). We used the five topics as a conceptual framework because they capture registry-level practices with clear ethical relevance for potential participants’ decision. Each topic was addressed in a separate set of questions in our interview guide. In our questions, however, we used the terms ‘funding’ instead of ‘conflicts of interest’, and ‘data handling’ instead of both ‘informed consent’ and ‘privacy and data protection’ because the original phrasing in Van den Akker et al. (2024) may not have been intuitive to participants in this study. ‘Funding’ represents what we considered to be the most pronounced conflict of interest for patient registries and the term ‘data handling’ was sufficiently broad that we could follow up with more specific questions. Besides asking about *what* information participants find useful, we asked *how* they would prefer to see that information communicated. To provide context for the participants’ responses we also asked who they would prefer to have (and not have) access to registry data, and about possible rewards for participating in registries. Figure 1 illustrates how the initial deductive ethics categories were refined into final categories and subcategories during analysis.

**Figure 1.**
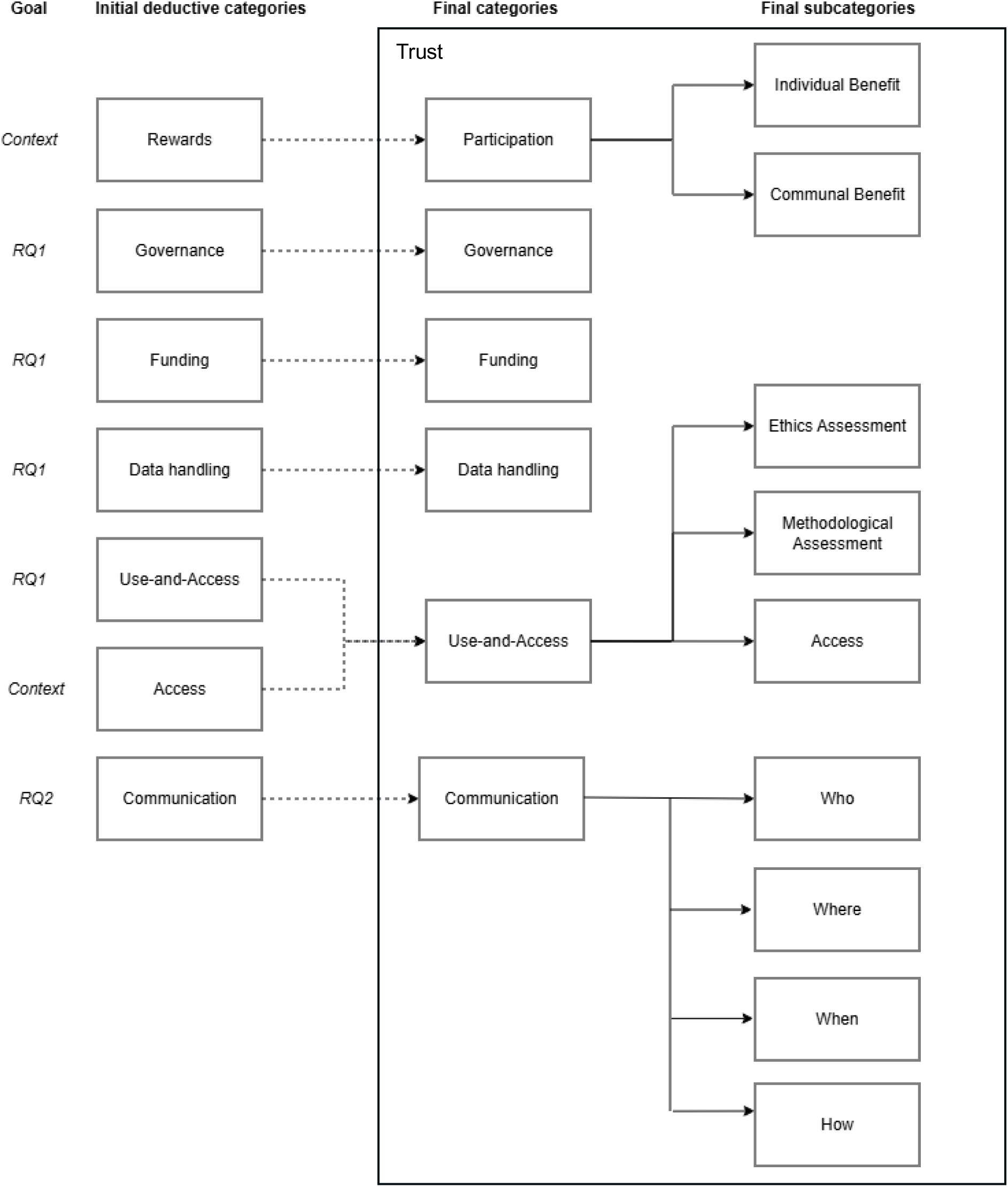
The hierarchical coding structure. *Note*. Trust was coded as a cross-cutting analytic concept that could co-occur with any category/subcategory. RQ = Research question.

Based on a pilot interview (with a patient representative with another diagnosis than MS so not included in the data analysis) and throughout the data collection we made slight modifications to specific interview questions, most of them to provide more clarity to the interviewee on what we meant by our questions. The full interview guide including a log with modifications we made throughout the interviews can be found at https://osf.io/e8p9w.

Interviews were conducted until data saturation (i.e., the degree to which new data repeat what was expressed in previous data; Saunders et al., 2018) was reached, which occurred when all three of the interviewers agreed that it was unlikely to encounter new information through additional interviews. Interviews took place from July 2024 until December 2024. After all interviews were conducted, we converted the recorded video files into audio files using VLC Media Player 3.0.21, and we transcribed the audio files verbatim via Transcription Stream, a locally run application based on OpenAI’s Whisper (Radford et al., 2023).

### Data analysis

The interviews were analyzed via directed qualitative content analysis (Hsieh and Shannon, 2005). The categories that functioned as the starting points for coding were the predefined topics from the interview guide (i.e., governance, funding, data handling, and use-and-access procedures), where the working definitions of these categories were developed and refined throughout the analysis. These four categories were intended to answer RQ1. To answer RQ2 we added the category ‘communication’, indicating how participants would want to be informed. Finally, we added two categories based on our contextual questions: ‘access’ to denote who participants think should have access to registry data, and ‘rewards’ to denote what participants think about any rewards for participating in a registry. We thus started coding with seven deductive categories (see the left-hand side of Figure 1) and assigned all relevant interview passages to one or more of these categories. As part of a coding pilot, two authors coded two interviews independently and discussed any inconsistencies together. After they were on the same page about the granularity and comprehensiveness of coding, tha first author coded all other interviews.

During coding, the predetermined categories were refined and inductive subcategories were developed to capture heterogeneity in the data. First, we downgraded the main category ‘access’ to a subcategory of ‘use-and-access’ because the question about who should have access to registry data typically came up when discussing use-and-access procedures. Second, we added the subcategories ‘ethics assessment’ (for topics related to the ethical review of research proposals sent to a registry) and ‘methodological assessment’ (for topics related to the methodological review of research proposal sent to a registry) to ‘use-and-access’ because we identified these as two types of assessment important in use-and-access procedures. Third, we relabeled the category ‘rewards’ into ‘participation’ and developed the subcategories ‘individual benefit’, and ‘communal benefit’ to capture different benefits participants (want to) see when sharing their data with a registry. Fourth, we developed the subcategories ‘who’, ‘where’, ‘when’, and ‘how’ to the category ‘communication’ to specify in more detail participants’ preferences for receiving information about the registry. Figure 1 provides an overview of the categories and subcategories we developed before and during data analysis. The data material was coded at both category and subcategory levels, where applicable.

To find more fine-grained meaning than the categories and subcategories allowed, we developed at least one inductive code for each of the text passages within a category, where separate codes were given to passages that involved approximately the same meaning or topic. Codes were thus more meaningful and lower-level descriptions of a (sub)category. For example, the category ‘communication’ includes the codes ‘not via website because untrustworthy’ and ‘after diagnosis’ signifying some participants’ preferences for the mode and timing of registry communication. Per (sub)category, we went over all the codes to establish underlying patterns in the data and to identify individually relevant content. The results section consists of a summary of these underlying patterns and individual pieces of content. An overview of all categories, subcategories, and codes can be found in our data file at https://osf.io/eknat. A post hoc secondary assessment of all (sub)categories to evaluate whether the codes matched the coded text passages was done for each final category. Any differential assessments were reconciled with the first author, after which the data file was updated. We used the ATLAS.ti software (version 25) for our coding. The quotations we extracted from the interviews were anonymized by redacting place names, specific organizations, personal information, and any other identifying words or phrases. Table 2 provides illustrative quotations for each of the categories and subcategories we identified.

**Table 1.** Demographics of the participants (N = 21).

| **Gender, n (%)** |  |
| --- | --- |
| Woman | 13 (61.9) |
| Man | 8 (38.1) |
| **Country, n (%)** |  |
| Belgium | 4 (19.0) |
| Croatia | 2 (9.5) |
| Denmark | 4 (19.0) |
| Germany | 2 (9.5) |
| Greece | 2 (9.5) |
| Netherlands | 2 (9.5) |
| Portugal | 1 (4.8) |
| Serbia | 2 (9.5) |
| Spain | 2 (9.5) |
| **Age, mean [min; max]** | 48.8 [31; 80] |
| **MS diagnosis, n (%)** |  |
| Yes | 20 (95.2) |
| No | 1 (4.8) |
| **MS duration, mean [min; max]** | 16.9 [3; 30] |
| **Patient representative, n (%)** |  |
| Yes | 14 (66.7) |
| No | 7 (33.3) |
| **Current or previous registry enrollment, n (%)** |  |
| Yes | 11 (52.4) |
| No | 7 (33.3) |
| Not sure | 3 (14.3) |

**Table 2.** Illustrative quotations for each of the categories and subcategories we identified.

| Category | Subcategory | Code | Quotation |
| --- | --- | --- | --- |
| Trust | NA | Trust comes from third parties | "I don't personally imagine wanting to wrap my head around that level of information regarding a registry. I would normally accept entry into registries which are already part of trusted institutions of some kind. For example, known universities or the public system. Especially as a person with a chronic illness, I wouldn't want to have to get that level of information. If I didn't have an initial feeling of trust, I just wouldn't go into it." (ID 16:8) |
| Participation | Individual benefit | General results | "And another thing that I think could be useful is also how would the communication be from the registry back to the patient. What is done with that information to show that it's not just sitting there for researchers to research, but there's actually an output - if that is possible, of course." (ID 11:7) |
| Participation | Communal benefit | Helping others | "I always say, for me, it wouldn't help anymore. But for the young women, for example, it will help." (ID 15:12) |
| Governance | NA | Patient representation | "Patient representatives should be in this executive board." (ID 7:9) |
| Funding | NA | Commercial vs. government | "For example, using the word: "This is a non-commercial entity that's funded by the governments of the European Union.", I mean, if I hear something like that and I think: "Okay, this is neutral, this is okay". If you say: "It's a pharmaceutical company", then I'm becoming a bit wary. I may in certain situations still say "It's fine", I wouldn't really be much more critical then. So, I think that's the kind of information you need to know. If it's pharmaceutical or commercial, then you need to know how is the safeguard. It's never completely safeguarded when it's commercial. But, I think then you want to know that. But if it's government and it's an independent institution, then I think/ well, I tend to trust these things." (ID 9:17) |
| Data handling | NA | Not too much detail | "Don't go into too much detail there because that will just frighten people. And then you say: "Oh, GDPR". People start thinking of everything that can go wrong. And they hear things on the radio, or on TV, or something about Facebook stealing so many things or just opening up all the information they have about their customers. So, briefly, just say it's all protected, following the privacy rules in your country and worldwide: "We're only going to use it for research."" (ID 11:6) |
| Use-and-access | Ethics assessment | The agency of the patient should be | "Okay let's say roughly it has to protect what ethical decision making is, is to |
|  |  | protected | protect the patient, to protect the researcher, and to protect the whole community in such a way that each three of them have dignity, meaning the right to make their own decisions." (ID 6:14) |
| Use-and-access | Methodological assessment | Methodological assessment is important | "It should be possible for the registry to say, okay we keep the data, but we don't just give it out to everybody. If the methodology is not up to standard, for instance, or there are doubts about what's going to happen with the data, then I would say keep it safe and don't open to certain parties." (ID 11:28) |
| Use-and-access | Access | Pharma should have access | "I personally/ I think pharma is great. Many, many patients are worried and scared of pharma, that they do bad things with their data." (ID 11:20) |
| Communication | Who | Via medical staff because of trust relationship | "Oh [thinking]. Via the channels that patients trust. So, that's either their medical people or like the MS patient organizations, patient advocates. Yeah, but through people they trust" [ID 18:37] |
| Communication | Where | Not via a website because untrustworthy | "If it's on a website, then you can just change it without telling me. And maybe that's a good thing and it could also be a bad thing. So [laughing], yeah. It's easier for you if it's a website and it's easier for me in five years to check your website but then again it's not the agreement we had. So I would also prefer it on email." (ID 20:11) |
| Communication | When | After diagnosis | "At a later date after diagnosis, there should be a talk about registries. Maybe it can be mentioned in the first day about health data, but certainly later on when they can actually think straight. So I think that would be a first one." (ID 11:34) |
| Communication | How | Written first | "Yeah, I mean, the first step could be via email and then they could have the option to speak to somebody about it, if necessary. Because there's some people that might just say "No, I've understood everything and I'm fine with it and I don't need that." But there might be, particularly older people might need to have that extra contact." (ID 14:26) |

The study is reported in accordance with the Standards for Reporting Qualitative Research (SRQR, O’Brien et al., 2014). A checklist with page numbers for the different SRQR items can be found at https://osf.io/3yqhm.

## Results

In total, 40 patients or patient representatives agreed to participate in an interview and filled in the pre-interview screening form. We then gradually invited them over a period of six months. Once we had interviewed five participants from Denmark, we stopped scheduling interviews with additional individuals from that country to prevent the sample from becoming too homogeneous. We reached out to three people from Croatia after they showed initial interest, but we were not able to establish a meeting with them. Five other individuals from Croatia were not invited because we already reached data saturation. In the end, we conducted 22 interviews, typically lasting between 60 and 75 minutes, but one of those (a person from Denmark) was cut off about halfway through for unclear reasons. Because we were not able to ask all our questions and could not re-establish contact with the participant then or later, we discarded the data from that interview. As such, our analysis is based on interviews with 21 people. All but one were people with MS (one participant was a patient representative without the disease) and two-thirds were patient representatives. Table 1 provides an overview of participant demographics.

We continue the results section with a narrative description of our findings per category. We start with a section about trust. During coding, trust emerged as salient across multiple content categories rather than being confined to a single domain. As such, we treated trust as a cross-cutting analytic concept and coded it in parallel so that it could co-occur with any category or subcategory in Figure 1. The data file (https://osf.io/eknat) includes a column called ‘Trust’ that indicates the quotations we deemed relevant to the concept of trust. After that, we continue with a section about participation to introduce participants’ general thoughts about (not) participating in a registry, followed by sections highlighting our findings in the remaining categories. The data file with anonymized quotations, quote IDs, (sub)categories, and codes can be found at https://osf.io/eknat.

### The role of trust

Across the interviews, an important insight emerged: trust is key for people with MS to decide whether they want to participate in a registry, for their assessment of what information is important to guide their decision, and how they would like this information to be communicated. Interestingly, trust was often based on endorsements from third parties the participants already knew and trusted, like universities or government agencies. Reassurance from these recognized and reputable sources was seen as an effective way to bolster confidence in a registry, for example in terms of the likelihood of data leaks. Trust could also be derived from the person or organization providing the initial information about a registry and requesting that individuals share their data. Participants mentioned that they would be happy to share data if their physician or someone else from their medical team would recommend or ask them to, simply because it is a trustworthy figure. If a registry would communicate through a patient organization, participants would be similarly positive about sharing their data. Some participants even said that trustworthy endorsements or communicators were sufficient and that additional detailed information about a registry’s governance structures, funding, and data handling procedures were not necessary anymore.

When trust was not established via external affiliations or familiar connections, participants emphasized that information received from a registry would allow them to assess whether it is trustworthy. An important indicator of trust was clear and transparent information, for example about a registry’s business plan. With regard to funding information, one participant summarized the relationship between trust and transparency by stating that the less they knew about a registry’s affiliation, the more information they needed (Quote ID = 18:20). A particularly strong factor in building trust was the inclusion of patient representatives on a registry’s governing boards. However, it was emphasized that these representatives should be meaningful contributors, not merely symbolic figures.

### Participation

Across the board, participants believed that patient registries could be very effective in finding treatments or even the underlying cause of (the different variants of) MS. Participants expressed a strong communal motivation for registry participation: they want to contribute to improve MS treatments, even if they were aware that any benefits would not come in their own lifetime. One participant stressed that the country of origin could play a role in shaping such a sense of community because civic participation is encouraged more in some countries than others.

While *communal benefit* was most salient, some participants also saw potential *individual benefit* from participating in a registry. Participants outlined some situations in which monetary compensation might be seen as a personal benefit and could be desirable. Most importantly, this was the case if the disease limited their ability to work, causing financial strain, or when participation required logistical effort, such as traveling to another city. Finally, it was stated that if patient data were to be used for commercial purposes, financial compensation would make more sense. In such cases, participants emphasized the importance of setting the amount carefully to balance intrinsic and extrinsic motivation. However, concerns about undesirable incentives were also raised, mainly in the sense that monetary rewards could attract participants that provide lower quality data, for example in terms of desirable or unusable responses.

Personal health information itself was also seen as a type of individual benefit. Participants expressed a desire to know whether a registry would allow them to access their individual data. This access could help them track changes in their symptoms over time. That way, a registry could serve as a centralized place to track patients’ health data, possibly even allowing them to add personal inputs like diary entries. However, one participant cautioned that such access might be distressing or lead to excessive focus on personal health.

Additionally, participants were interested in whether the registry would provide general information about the disease or general results derived from the registry data, such as scientific publications. These broader insights would help them stay informed about the disease and its treatment and would give them a sense of gratification from sharing their data. However, one participant noted that giving individual patients access to general scientific results about MS could be dangerous because they do not always have the scientific education to appropriately interpret such results.

### What information is considered important

#### Governance

Participants held varied views about how much they wanted to know regarding the governance structure of registries. While some viewed governance transparency as critical to trust, others considered it too complex or unnecessary. Generally, participants wanted at least some information, such as the qualifications of the board members, the degree of independence of the board, and whether there were mechanisms in place to ensure accountability.

#### Funding

Regarding the funding of registries, some participants were very clear that they wanted information, particularly about the source of funding. The interviewees made clear that they would be more open to sharing their data if a registry was funded by an MS society, a university, or a public institution, but not if it was funded by commercial medical companies. Distrust towards medical firms originated from skepticism about their desire to improve the lives of patients, their capacity to keep data safe, and potential conflicts of interest.

Because of the importance placed on the source of funding, participants noted that they would like information about the commercial nature of the registries. However, most participants were not in favor of being provided with detailed financial information. Other participants emphasized the need for patients to be aware of how the money is distributed *within* the registry, for example to assess conflicts of interest.

#### Data handling

For data handling (i.e., privacy and data protection), the participants’ perspectives were polarized. Some participants were highly interested and wanted access to detailed documentation on how their data would be processed by the registry, including detailed information about procedures or links to regulatory frameworks. Others, however, were not interested in these details at all. A major reason for not being interested in specific information about privacy and data protection was that the participants trusted the people or institutions associated with the registry. For some, the registry merely mentioning that they comply with privacy regulations like the General Data Protection Regulation (GDPR) was sufficient, especially if they already had a high level of trust towards the registry or when the punishment for non-adherence would be high. Others wanted a guarantee that their data would be anonymous, or at least some information about the degree of anonymization. One participant proposed an audit or certification system for registries, where a simple visual checkmark would indicate compliance with regulations (Quote ID = 10:10). This would allow patients to avoid sifting through complex legal or technical details. Finally, some participants wanted to know whether they would be able to opt out of participation.

Participants who wanted information from the registry about data handling typically emphasized that they would like to know *who* would have access to the data and *where* it would be going, *what* purposes their data would be used for (i.e., the aim of the registry or the researchers using the data), and *how* their data would be used (i.e., whether it would be handled anonymously and/or securely). Some participants also wanted to know what specific data they would need to provide. Interestingly, a few participants reported that their confidence in a registry would actually increase if a data breach occurred and was disclosed publicly. For them, transparency, even in the face of a security incident, was a sign of integrity and accountability.

#### Use-and-access

In general, participants believed that potential data users like researchers should provide detailed information to registries about themselves, and about the study’s design and goals, possibly with an added layer of verification by a university or other trusted entity. This is in line with worries voiced by participants about the resharing or (re-)selling of their data by data users. Participants were clear-cut in their opposition to such practices. To prevent data from being disclosed or sold without authorization, some interviewees argued that data access should be restricted to anonymized datasets or to the local digital environment of the registry itself. Others stated that it should be very clear in agreements between the registry and data users that the data would only be used for a specific purpose, and that clear punishments could help reinforce such agreements. There was substantial heterogeneity in opinions about who should have access to data from patient registries. Some participants raised concerns about the trustworthiness of pharmaceutical companies, while others did not take much issue with them having access or thought that patients do not mind just because they are ignorant about the ramifications of sharing their data.

When asked whether participants would want to know whether a registry’s use-and-access procedure includes their own *ethics assessment* or *methodological assessments* of proposed research studies, some participants seemed to not fully understand what such assessments would entail while others expressed uncertainty about what they preferred. Many participants did support the idea of some sort of oversight. Some participants mentioned that it would be better if an external party would do ethical or methodological checks because registries themselves often lack the resources or expertise for such checks. Participants generally found ethical checks more important than methodological checks, although a few participants pointed out that a basic methodological check could still be useful to determine whether a proposed study is feasible and appropriate based on the registry’s available data.

### How information should be communicated

#### Communication

*How* information about registries should be communicated to people with MS involves knowing *who* should communicate, *when* to communicate, *where* to find the information, and *how* to communicate.

Regarding *who* should communicate about a registry, some participants preferred to receive information from their doctor or someone else from the medical staff, citing established trust and familiarity. However, others raised concerns about this approach. Not all patients have a positive relationship with their healthcare provider, and one participant felt that the involvement of the medical staff could even create pressure to participate, making it harder to decline (Quote ID = 8:21). Time constraints in clinical settings were also mentioned when discussing the complexities of communication via doctors, with participants noting that doctors may not be able to take on additional responsibilities related to registry promotion.

In terms of *when* to contact participants, several participants emphasized that introducing registry information immediately after diagnosis is not ideal, as patients are often overwhelmed and unable to absorb complex or unfamiliar content at that stage.

Regarding *where* and *how,* some participants preferred an initial face-to-face conversation followed by written materials. Others favored receiving written information first and then having the opportunity to ask questions in a personal meeting. This diversity in preferences reflects the heterogeneity in how patients want to be addressed, and some participants also mentioned this heterogeneity explicitly, for example by pointing out differences in cognitive or other abilities. In this context, they often mentioned that older people have more trouble with technological communication and should therefore be preferentially addressed in person.

That being said, digital or written information was seen as valuable because it generally allows people to read back the information at a later time, especially when simple language is used. Participants also proposed more creative communication methods, including awareness campaigns, information videos, and podcasts. One participant noted that visual presentation, for example via diagrams, could be helpful alternatives to text-heavy materials (Quote ID = 11:9). Skepticism was, however, expressed toward information found on websites, as participants felt such content could be unilaterally changed while still appearing official.

Balanced communication of information appears to be key. This became particularly clear throughout the interviews, for example, when it came to information on data protection and privacy in informed consent forms. Many participants felt overwhelmed by the volume and complexity of information presented in informed consent forms. Some admitted not reading the forms in full, particularly when the cognitive symptoms of MS made processing of dense information more difficult. One participant suggested that oral explanations alongside written consent forms could help ensure patients fully understand what they are agreeing to (Quote ID = 3:23).

## Discussion

Through semi-structured interviews with patients and patient representatives, this study aimed to identify what specific information people with MS consider important to make informed decisions about participating in patient registries (RQ1), and how they would prefer this information to be communicated (RQ2).

A central finding was that trust functioned as a cross-cutting factor, shaping how participants evaluated registry practices, what information they deemed sufficient, and how they wanted that information to be delivered. The essential role of trust in sharing data has previously been shown for people using health data portals in Croatia and New Zealand (Bosanac and Stevanovic, 2022; Dobson et al., 2023), the general population in Singapore (Lysaght et al., 2021), people with chronic conditions in the United States (Lee et al., 2016), and in a systematic review of the literature (Howe et al., 2018). The current study adds nuance by showing that trust interacts with the level of detail of the provided information. For many participants, trust in the institutions or individuals associated with a registry reduced the need for detailed information. Hence, trust in associated parties functions as a sort of heuristic of the trustworthiness of a registry, where recognition and reputation are likely to play important roles. In the context of data sharing, this mechanism was found previously by Hermansen et al. (2024) for people with cancer, and by Goodman et al. (2025) for a representative sample of the general population in the United Kingdom. Where trust was not established enough, participants emphasized the importance of transparency and detailed explanations to enable their own assessment of the registry’s trustworthiness.

Across the interviews, participants emphasized the need for clear information on who will access their data and how the data will be used, which is in line with the results of earlier studies (Cascini et al., 2024; Dobson et al., 2023). Many also wanted to know which data they would be asked to provide, whether this data would be anonymously processed, and whether they could access individual-level data or summary results in return. Access to individual-level data was often seen as a reward for data sharing and has previously also been associated with feelings of respect and the perception of control (Dobson et al., 2023). Our findings are also in line with earlier studies showing that MS patients (Dennison et al., 2018) and patients with other chronic conditions (Council of Medical Specialty Societies, 2019; Lee et al., 2016) have a high need for information about their specific diagnostic situation and their disease in general. Indeed, the Agency for Healthcare Research and Quality (2020) emphasizes that dissemination of registry research results to registry participants should be formally included in data transfer agreements of registries. This should help combat ‘helicopter research’, where participants feel that researchers collect their data and then never follow up or provide any feedback.

Interestingly, few participants wanted to examine technical data protection measures in detail. A statement that the registry complies with established standards (e.g., GDPR) was often deemed sufficient. These findings seem to contrast with previous studies that show that privacy and data protection are key concerns for patients (Baines et al., 2024; Dobson et al., 2023). Indeed, these prior studies indicated that trust comes from clear and transparent communication about privacy and data protection details, while our study indicates a reversed relationship where an initial level of trust leads to less desire for such details. In our study, participants stated that they were sometimes overwhelmed by the amount of information provided in consent forms, which is consistent with studies showing low comprehension of such forms (Manta et al., 2021; Tam et al., 2015). Interestingly, shorter recruitment letters have been shown previously to increase enrollment in randomized controlled trials (Johansen et al., 2025), a finding which may translate to the context of patient registry participation.

Preferences regarding how information should be communicated varied strongly but there was widespread agreement that personal contact through trusted intermediaries was often preferred, although the usefulness of written information was also noted. Registries would do well to take the context of the specific disease into account and accommodate diverse information needs and cognitive capacities, potentially using a layered (e.g., providing only summary information but with links to more detail) or multimodal approach (e.g., communicating face-to-face but also using video messages and a digital newsletter). A preference for a layered, multimodal approach was also found for users at patient portals (Reynolds et al., 2021) and is recommended by the Agency for Healthcare Research and Quality (2020).

One strength of this study is the diverse sample, with participants from different European regions, of various age groups, and with different disease durations. Nevertheless, the sample primarily consisted of patient representatives with MS, whose perspectives may differ from those of the broader MS population. Representatives may be more accustomed to engaging with institutional stakeholders and may therefore emphasize collective or governance-related considerations more strongly than individual experiential concerns (see Learmonth et al., 2009; Van de Bovenkamp and Trappenburg, 2011). In addition, the interviews focused more heavily on attitudes than on actual behavior, which may not always translate into decision-making in practice. For example, it is unclear whether the likelihood of patients participating in a registry indeed increases when they are provided with access to individual-level data or when affiliated institutions are presented transparently.

Other limitations speak to the study’s procedures. First, only one coder did the main analysis. We chose this pragmatic approach because our coding pilot mainly showed discrepancies in the granularity of coding (i.e., the amount of detail captured in the quotes and synthesized in the codes), not in the categorizations themselves. To help mitigate the potential bias of the single coder approach, a secondary assessment was conducted where all the codes were inspected by another member of the team, leading to some changes in the (sub)categories and codes. To enhance transparency, we openly provide the coding tree with illustrative quotations. This allows readers to directly look up the quotes for each specific result themselves.

Finally, while this study focused specifically on MS, there is no strong theoretical reason to expect that the information priorities identified in this study would substantially differ when it comes to other chronic conditions. However, this is an empirical question and comparative studies across disease contexts would be informative.

To conclude, this study offers practical insights to registry owners and other stakeholders into what information people with MS consider important when deciding to contribute data to patient registries, and how they prefer this information to be communicated. By applying an ethics-informed framework to the perspectives of people with MS on information needs and communication preferences for decision-making about registry participation, the study shows that registry practices such as governance, funding, data handling, and use-and-access procedures are not only institutional or technical matters, but also shape patients’ assessments of trustworthiness and participation. For the participants, building and maintaining trust, through credible governance, meaningful patient involvement, transparent communication, and clear feedback about the value of contributed data, appears central to informed decision-making regarding registry participation. Registries should adopt flexible, layered communication strategies that acknowledge varying levels of trust and differing preferences and capabilities for detail and modalities.

## Artificial intelligence statement

ChatGPT was used to improve the language, clarity, and structure of the text, to assist with restructuring data files, and to correct the formatting and consistency of references. The AI tool was not used to generate research findings.

## Acknowledgments

The authors wish to thank all the participants of the interviews for the time and effort they took to provide their valuable perspectives. We would also like to thank Daniel Strech for his methodological input, Ingo Przesdzing for his help with the transcription software, Mohsharif Nasrulloeva for her help with inviting participants, and Merle-Marie Pittelkow for feedback on a first draft of the manuscript.

## Author contributions

OvdA, SdV, DA, and FW conceived and designed the study. OvdA conducted interviews together with either FW or DA, and coded the interviews with methodological support from SS. OvdA also curated the data file and wrote the first draft of the manuscript. All authors contributed to the development of the interview guide. OvdA and FW conducted the coding pilot and discussed coding procedures. SS, SdV, DA, and FW conducted secondary assessments of assigned coding categories. FW, DA, SdV, and SS provided feedback on the first draft. All authors contributed to the interpretation of the findings, critically reviewed and revised the manuscript, and approved the final version.

## Statements and Declarations

### Ethical considerations

This study was approved by the Ethics Committee of Charité - Universitätsmedizin Berlin (EA1/137/24). The study was completed in line with the principles of the Declaration of Helsinki.

### Consent to participate

Informed consent for conducting the interviews was requested and received from all participants.

### Consent for publication

Written informed consent for publication of anonymized data and quotations was obtained from all participants. All potentially identifying information has been removed.

## Data availability

The materials used in this study, and the data used for our analysis (the coding tree and relevant quotations from the interviews) can be found at https://osf.io/jwnqr.

## Positionality statement

The itnerviews were typically conducted by OvdA with either FW or DA present as back-up to ask follow-up questions. OvdA is a meta-researcher with a focus on the ethics and transparency of studies using existing data. FW and DA are medical researchers with a focus on using real-world data for drug regulation and pharmacoepidemiology.

## References

Agency for Healthcare Research and Quality (2020) Registries for Evaluating Patient Outcomes: A User’s Guide. 4th ed.

Baird W, Jackson R, Ford H, et al. (2009) Holding personal information in a disease-specific register: The perspectives of people with multiple sclerosis and professionals on consent and access. Journal of Medical Ethics 35(2): 92–96.

Baines R, Stevens S, Austin D, et al. (2024) Patient and public willingness to share personal health data for third-party or secondary uses: Systematic review. Journal of Medical Internet Research 26: e50421.

Bosanac D and Stevanovic A (2022) Trust in E-health system and willingness to share personal health data. In: Informatics and Technology in Clinical Care and Public Health, pp. 256–259. IOS Press.

Cascini F, Pantovic A, Al-Ajlouni YA, et al. (2024) Health data sharing attitudes towards primary and secondary use of data: A systematic review. eClinicalMedicine 71.

Council of Medical Specialty Societies (2019) Engaging Patients in Clinical Registries. Available at: https://cmss.org/wp-content/uploads/2020/03/Key-Concepts-in-Patient-Engagement-1.pdf (accessed 19 July 2026).

Dennison L, Brown M, Kirby S, et al. (2018) Do people with multiple sclerosis want to know their prognosis? A UK nationwide study. PLOS One 13(2): e0193407.

Dobson R, Wihongi H and Whittaker R (2023) Exploring patient perspectives on the secondary use of their personal health information: An interview study. BMC Medical Informatics and Decision Making 23(1): 66.

European Medicines Agency (2021) Guideline on Registry-Based Studies (EMA/426390/2021), 22 October. Available at: https://www.ema.europa.eu/en/documents/scientific-guideline/guideline-registry-based-studies_en.pdf (accessed 19 July 2026).

European Network for Health Technology Assessment (EUnetHTA) (2019) Registry Evaluation and Quality Standards Tool (REQueST). Available at: https://catalogues.ema.europa.eu/system/files/2025-02/05.01.0301%20Feasibility%20Documentation%20%20-%20Registry%20Evaluation%20and%20Quality%20Standards%20Tool%20%28REQueST%29%20_%2010-Sep-2023_Redacted.pdf (accessed 19 July 2026).

Flachenecker P and Stuke K (2008) National MS registries. Journal of Neurology 255: 102–108.

Goodman JR, Costa A and Milne R (2025) Trust and perceived trustworthiness in health-related data sharing among UK adults: Cross-sectional survey. Journal of Medical Internet Research 27: e83533.

Hageman IC, Van Rooij IA, De Blaauw I, et al. (2023) A systematic overview of rare disease patient registries: Challenges in design, quality management, and maintenance. Orphanet Journal of Rare Diseases 18(1): 106.

Hermansen A, Pollard S, McGrail K, et al. (2024) Heuristics identified in health data-sharing preferences of patients with cancer: Qualitative focus group study. Journal of Medical Internet Research 26: e63155.

Hillert J and Stawiarz L (2015) The Swedish MS registry-clinical support tool and scientific resource. Acta Neurologica Scandinavica 132: 11–19.

Howe N, Giles E, Newbury-Birch D, et al. (2018) Systematic review of participants’ attitudes towards data sharing: A thematic synthesis. Journal of Health Services Research & Policy 23(2): 123–133.

Hsieh H-F and Shannon SE (2005) Three approaches to qualitative content analysis. Qualitative Health Research 15(9): 1277–1288.

Johansen ND, Preiss D, Mafham M, et al. (2025) Randomized experiment of recruitment letter design to maximize clinical trial enrollment. JAMA, 334(7): 633–635.

Jonker CJ, Bakker E, Kurz X, et al. (2022) Contribution of patient registries to regulatory decision making on rare diseases medicinal products in Europe. Frontiers in Pharmacology 13: 924648.

Kalkman S, Mostert M, Gerlinger C, et al. (2019) Responsible data sharing in international health research: A systematic review of principles and norms. BMC Medical Ethics 20(1): 21.

Kalkman S, Van Delden J, Banerjee A, et al. (2022) Patients’ and public views and attitudes towards the sharing of health data for research: A narrative review of the empirical evidence. Journal of Medical Ethics 48(1): 3–13.

Learmonth M, Martin GP and Warwick P (2009) Ordinary and effective: The Catch-22 in managing the public voice in health care? Health Expectations 12(1): 106–115.

Lee SB, Zak A, Iversen MD, et al. (2016) Participation in clinical research registries: A focus group study examining views from patients with arthritis and other chronic illnesses. Arthritis Care & Research 68: 974–980.

Lysaght T, Ballantyne A, Toh HJ, et al. (2021) Trust and trade-offs in sharing data for precision medicine: A national survey of Singapore. Journal of Personalized Medicine 11(9): 921.

Manta CJ, Ortiz J, Moulton BW, et al. (2021) From the patient perspective, consent forms fall short of providing information to guide decision making. Journal of Patient Safety 17(3): e149–e154.

O’Brien BC, Harris IB, Beckman TJ, et al. (2014) Standards for reporting qualitative research: A synthesis of recommendations. Academic Medicine 89(9): 1245–1251.

Radford A, Kim JW, Xu T, et al. (2023) Robust speech recognition via large-scale weak supervision. In: Krause A, Brunskill E, Cho K, et al. (eds) Proceedings of the 40th International Conference on Machine Learning, pp. 28492–28518.

Reynolds TL, Ali N and Zheng K (2021) What do patients and caregivers want? A systematic review of user suggestions to improve patient portals. In: AMIA Annual Symposium Proceedings 2020: 1070.

Saunders B, Sim J, Kingstone T, et al. (2018) Saturation in qualitative research: Exploring its conceptualization and operationalization. Quality & Quantity 52(4): 1893–1907.

Simon SR, Evans JS, Benjamin A, et al. (2009) Patients’ attitudes toward electronic health information exchange: Qualitative study. Journal of Medical Internet Research 11(3): e30.

Tam NT, Huy NT, Thoa LTB, et al. (2015) Participants’ understanding of informed consent in clinical trials over three decades: Systematic review and meta-analysis. Bulletin of the World Health Organization 93: 186–198H.

Van de Bovenkamp HM and Trappenburg MJ (2011) Government influence on patient organizations. Health Care Analysis 19(4): 329–351.

Van den Akker OR, Stark S and Strech D (2024) Ethics practices associated with reusing health data: An assessment of patient registries. BMC Medicine 22(1): 1–10.

Whiddett R, Hunter I, Engelbrecht J and Handy J (2006) Patients’ attitudes towards sharing their health information. International Journal of Medical Informatics 75(7): 530–541.

